# Sensitive detection of SARS-CoV-2-specific-antibodies in dried blood spot samples

**DOI:** 10.1101/2020.07.01.20144295

**Authors:** Gabriella L. Morley, Stephen Taylor, Sian Jossi, Marisol Perez-Toledo, Sian E. Faustini, Edith Marcial-Juarez, Adrian M. Shields, Margaret Goodall, Joel D. Allen, Yasunori Watanabe, Maddy L. Newby, Max Crispin, Mark T. Drayson, Adam F. Cunningham, Alex G. Richter, Matthew K. O’Shea

## Abstract

**Importance:** Population-wide serological testing is an essential component in understanding the COVID-19 pandemic. The logistical challenges of undertaking widespread serological testing could be eased through use of a reliable dried blood spot (DBS) sampling method.

**Objective:** To validate the use of dried blood spot sampling for the detection of SARS-CoV-2-specific antibodies.

**Design, setting and participants:** Eighty-seven matched DBS and serum samples were obtained from eighty individuals, including thirty-one who were previously PCR-positive for SARS-CoV-2. DBS eluates and sera were used in an ELISA to detect antibodies to the viral spike protein.

**Results:** Specific anti-SARS-Cov-2 spike glycoprotein antibodies were detectable in both serum and DBS eluate and there was a significant correlation between the antibody levels detected in matched samples (r = 0.96, p<0.0001). Using serum as the gold standard in the assay, matched DBS samples achieved a Cohen’s kappa coefficient of 0.975 (near-perfect agreement), a sensitivity of 98.1% and specificity of 100%, for detecting anti-spike glycoprotein antibodies.

**Conclusions and relevance:** Eluates from DBS samples are a reliable and reproducible source of antibodies to be used for the detection of SARS-CoV-2-specific antibodies. The use of DBS sampling could complement the use of venepuncture in the immunosurveillance of COVID-19 in both low and high income settings.

## Introduction

Confirmation of a diagnosis for acute coronavirus disease 2019 (COVID-19) is dependent upon the detection of RNA from the causative pathogen, severe acute respiratory syndrome coronavirus 2 (SARS-CoV-2). In contrast, while serology is less useful for diagnosing acute stages of infection in most individuals, it can help diagnose atypical presentations of SARS-CoV-2 and asymptomatic infection^1^. Furthermore, serology is valuable for determining prior viral exposure at a population level, allowing for a more comprehensive understanding of the epidemiology of SARS-CoV-2, particularly given the limitations of polymerase chain reaction testing^2^. Antibody assessments can also help to establish transmission patterns, assess longitudinal humoral responses and identify correlates of protection, all of which may have a significant impact on public health and social policies^3,4^.

Currently, antibody testing for SARS-CoV-2 uses serum or plasma collected by venepuncture. The use of such sampling in large-scale seroepidemiological studies is limited by logistical challenges, resources and costs, together with the risk of SARS-CoV-2 exposure from direct patient contact. In contrast, dried blood spot (DBS) sampling is simple, inexpensive^5^ and can be self-collected then sent by postal services to laboratories for processing^6^. It is a well-established method for detecting antibodies against a variety of infections^7,8^ and antibodies collected on DBS cards are stable for prolonged periods^9^. Moreover, DBS sampling provides one solution to widening access to serological platforms in low- and middle-income countries (LMICs). Nevertheless, the potential role of the DBS to study SARS-CoV-2 seroprevalence has not been fully explored; and there is limited understanding of how to recover antibody from DBS. Here, we describe the validation of DBS samples against matched serum in a highly sensitive and specific SARS-CoV-2 Enzyme Linked Immunosorbent Assay (ELISA).

## Methods

### Participants and sample collection

Eighty-seven samples were collected from eighty volunteers at the University Hospitals Birmingham (UHB) NHS Foundation Trust (REC reference 2002/201) between 18/05/2020 and 03/06/2020. Five individuals provided duplicate samples and one individual provided triplicate matched samples. Three matched samples from SARS-CoV-2 antibody-negative volunteers (Clinical Immunology Service Reference ERN_16-178) were included for analysis; and for refining negative thresholds, a further seventeen pre-August 2019 DBS samples (REC reference 2002/20, IRAS reference 132132, UHB project reference RRK4136). Participants were healthy at the point of sampling and thirty-one matched samples (35.6%) were collected from PCR-positive subjects, on average 54±17 days from reported symptom onset. Participants were anonymised and SARS-CoV-2 PCR status was recorded as either positive, negative or unknown.

For DBS collection, finger-prick capillary blood samples (approximately 50 µL per spot) were collected onto forensic 226 grade DBS collection cards (Ahlstrom Munksjo, Germany) provided as part of the TakeATestUK postal testing kits^6^ (Saving Lives, UK), using standard methods^10^. DBS cards were stored at room temperature (RT) in individual sample bags with desiccant. Concomitantly, venous blood was collected from volunteers and serum was separated by centrifugation at 9,700 × *g* for 5 minutes, RT. Laboratory analysis was blinded to PCR status and SARS-CoV-2-specific-antibody results were reported as positive, negative or equivocal.

### DBS elution

To elute antibody from DBS cards, individual 12 mm diameter pre-perforated DBS spots were isolated using a sterile pipette tip and placed into a universal tube at a ratio of one spot to 250 µL 0.05% PBS Tween 20 (PBS-T). Tubes were briefly vortexed and incubated overnight at RT. DBS eluate was subsequently harvested into a microtube and centrifuged at 10,600 × *g* for 10 minutes at RT and stored at 4°C until use. Total IgG, IgA and IgM immunoglobulin concentrations of matched serum and DBS, plus pre-August 2019 DBS samples, was quantified by nephelometry using the automated COBAS 6000 (Roche, UK).

### SARS-CoV-2 ELISA

An ELISA to measure IgG, IgA and IgM against soluble, stabilised, trimeric SARS-CoV-2 spike (S) glycoprotein^11,12^, was performed as previously described^1,13^. Briefly, 96-well plates (Nunc, UK) were coated with 50 µL of 2 µg/mL spike (S) glycoprotein. Plates were blocked and sample diluted with 2% BSA PBS-T 0.1% (starting dilutions: 1:3 for DBS eluate and 1:15 for serum with 3-fold serial dilutions; or single dilutions of 1:10 for DBS eluate and 1:100 for serum). Mouse monoclonal anti-human HRP-conjugated antibodies (anti-IgG clone R-10 1:8000, anti-IgA MG4.156 1:4000 and anti-IgM AF6 1:2000; clones were generated at the University of Birmingham, available from Abingdon Health, UK) were diluted in 0.1% PBS-T. Plates were developed with TMB core (Bio-rad, UK) and stopped after 5 minutes with 0.2M H_2_SO_4_. Optical densities were recorded at 450 nm (OD_450_) using the Dynex Revelation automated liquid handler. Results were reported as SARS-CoV-2-anti-S antibody positive, negative or equivocal. The cut-off for negativity was less than the highest negative control; and for positivity, the mean of the negative controls plus 3 standard deviations (+3SD); between this range was equivocal.

### Statistical analysis

Statistical analyses were performed using Prism (version 8, GraphPad, USA). Correlations between continuous data were assessed using Spearman’s rank test and a p value <0.05 was considered statistically significant. DBS sample ELISA performance was assessed by calculating the sensitivity, specificity, positive and negative predictive values (PPV and NPV, respectively) with 95□% confidence intervals (CIs). The agreement between DBS and serum ELISA results was assessed by determining the Cohen’s kappa coefficient and Bland-Altman mean difference.

## Results

### SARS-CoV-2-anti-spike glycoprotein antibodies can be eluted from DBS samples

Quantification of total immunoglobulin concentrations in serum and DBS eluate was performed. A 7-11-fold reduction in mean immunoglobulin concentration (IgG, IgA and IgM) was observed in DBS eluate compared to matched serum (Fig 1a). To detect antibodies to SARS-CoV-2-anti-spike (S) glycoprotein, matched serum and DBS titration curves were generated. Specific anti-S antibodies were detectable in both serum and DBS eluate with higher responses observed in PCR-positive matched samples, whilst pre-August 2019 DBS samples were negative across all dilutions (Fig 1b and 1c).

**Figure 1.**
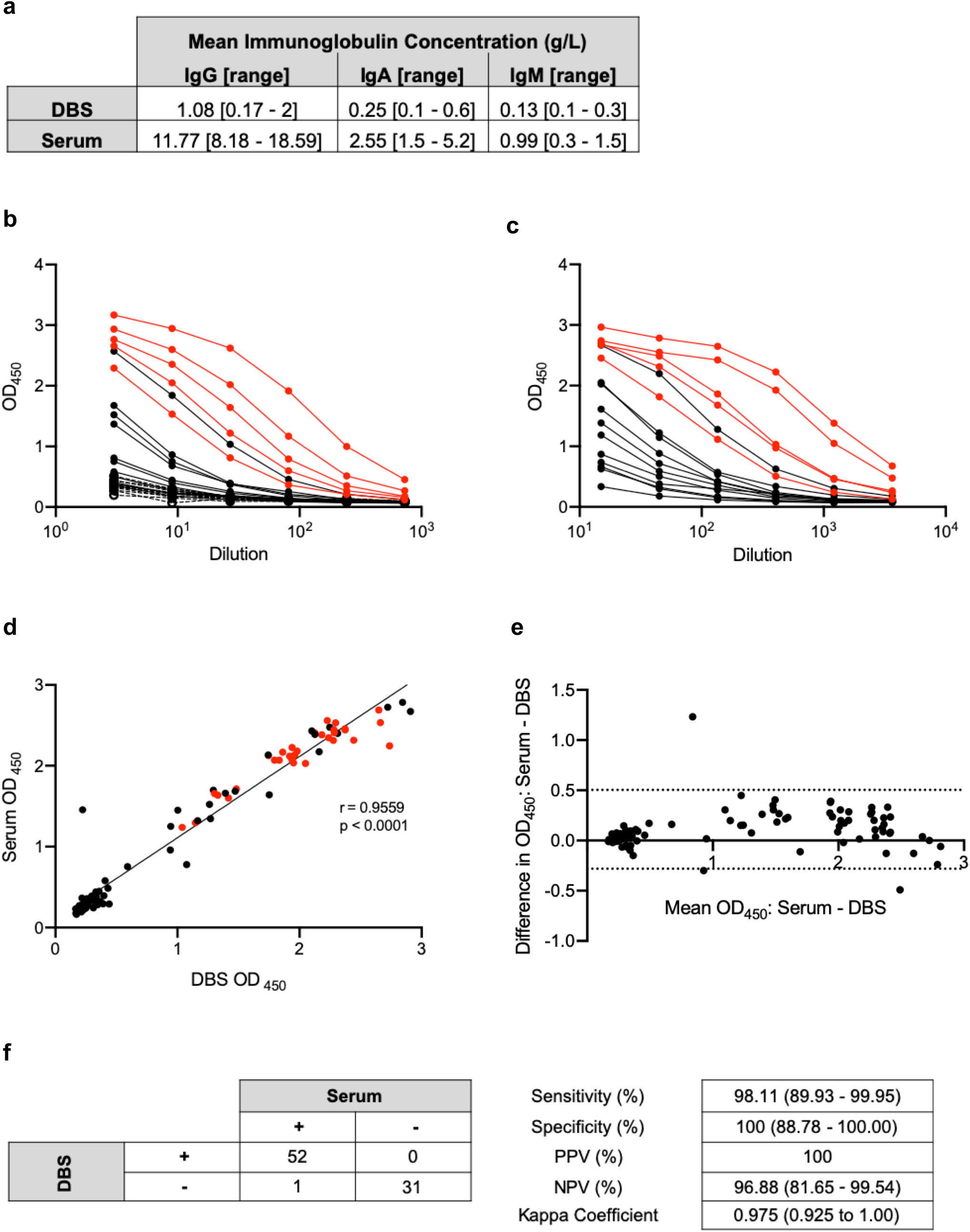
DBS sampling is effective for SARS-CoV-2 anti-S glycoprotein detection. **a**, Mean [range] concentrations of IgG, IgA and IgM measured in matched DBS eluate and serum samples (n=10 matched DBS and serum; n=5 pre-August 2019 DBS); **b**, DBS eluate titrations, in three-fold, with initial 1:3 dilution and **c**, serum titrations, in three-fold, with initial 1:15 dilution (red circle: PCR-positive (n=5); black circle: PCR-unknown (n=11); dashed line: pre-August 2019 DBS sample (n=11)); **d**, Correlation between DBS eluate (1:10) and serum (1:100) OD_450_ ELISA results (red circle: PCR-positive (n=31); black circle: PCR-unknown (n=56)); **e**, Bland-Altman mean difference comparison of DBS eluate (1:10) and serum (1:100) OD_450_ ELISA results (dashed lines represent 95% limits of agreement (−0.281 – 0.504)); **f**, 4×4 table of DBS eluate ELISA sensitivity and specificity (n=84 matched DBS and serum samples either positive or negative for the detection of SARS-CoV-2 anti-S glycoprotein; equivocal results (n=3) were excluded).

### Responses between matched serum and DBS samples correlate strongly

The OD_450_ detected by ELISA for matched DBS samples and sera, diluted 1:10 and 1:100 respectively, were plotted. There was a significant correlation between matched serum and DBS samples (r = 0.96 (95% CI: 0.93-0.97), p<0.0001) (Fig 1d) and minimal differences in results using either antibody source in the assay (Bland-Altman bias 0.11±0.20) (Fig 1e). Discordance occurred between only one matched sample, giving a Cohen’s kappa coefficient of 0.975. DBS samples achieved a sensitivity of 98.11% and specificity of 100% for detecting anti-S glycoprotein antibodies, when compared to serum (Fig 1f), with 100% of the PCR-positive samples (n=31) also antibody-positive in DBS eluate.

## Discussion

We show that DBS samples can be used for the detection of SARS-CoV-2-specific antibodies with high levels of sensitivity and specificity and compared well with matched serum samples. Taken together, these results demonstrate that DBS sampling could complement venepuncture for serological assessments, such as seroprevalence studies, during the COVID-19 pandemic.

A current limitation in antibody assays is the necessity for venepuncture by skilled phlebotomists. The use of DBS overcomes this limitation and introduces the opportunity for wider population level sampling and improved surveillance in those groups shielding from the infection. For example, DBS could be delivered using postal services^6^ to patients with chronic conditions, the immunocompromised and the elderly; groups which have been disproportionately affected by COVID-19^14^. Furthermore, the DBS method is simple and inexpensive^5,6^ which could enhance sampling in LMICs, amongst groups where venepuncture is culturally unacceptable or, in a geographically dispersed populous^15^.

## Data Availability

The authors confirm that the data supporting the findings of this work are available within the article.

## Acknowledgements

We would like to thank the University of Birmingham Clinical Immunology Service for their invaluable support in sample collection and processing; and thank Cynthia D’Aguilar and Julie Williams, from University Hospitals Birmingham (UHB) NHS Foundation Trust, for logistical support in sample collection. This work was supported the Wellcome Trust and was also supported by the National Institute for Health Research (NIHR) Birmingham Biomedical Research Centre at the UHB NHS Foundation Trust and the University of Birmingham. The views expressed are those of the author(s) and not necessarily those of the NIHR or the Department of Health and Social Care. This project was supported by the Saving Lives Charity (UK Charity Commission number 1144855) who kindly provided the TakeATestUK Dried Blood Spot postal collection kits. The work, in Prof. Max Crispin’s laboratory, was funded by the International AIDS Vaccine Initiative, Bill and Melinda Gates Foundation through the Collaboration for AIDS Vaccine Discovery (OPP1196345/INV-008813, OPP1084519 and OPP1115782), the Scripps Consortium for HIV Vaccine Development (CHAVD) (NIH: National Institute for Allergy and Infectious Diseases AI144462), and the University of Southampton Coronavirus Response Fund.

## References

1. Perez-Toledo M, Faustini SE, Jossi SE, et al. Serology confirms SARS-CoV-2 infection in PCR-negative children presenting with Paediatric Inflammatory Multi-System Syndrome. medRxiv. June 2020. doi:10.1101/2020.06.05.20123117

2. Wang W, Xu Y, Gao R, et al. Detection of SARS-CoV-2 in Different Types of Clinical Specimens. JAMA. March 2020.

3. Shields AM, Faustini SE, Perez-Toledo M, et al. SARS-CoV-2 seroconversion in health care workers. medRxiv. May 2020. doi:10.1101/2020.05.18.20105197

4. Long Q-X, Tang X-J, Shi Q-L, et al. Clinical and immunological assessment of asymptomatic SARS-CoV-2 infections. Nat Med. June 2020.

5. McDade TW, Williams S, Snodgrass JJ. What a drop can do: dried blood spots as a minimally invasive method for integrating biomarkers into population-based research. Demography. 2007;44(4):899–925.

6. Page M, Atabani SF, Wood M, et al. Dried blood spot and mini-tube blood sample collection kits for postal HIV testing services: a comparative review of successes in a real-world setting. Sex Transm Infect. 2019;95(1):43–45.

7. Vázquez-Morón S, Ryan P, Ardizone-Jiménez B, et al. Evaluation of dried blood spot samples for screening of hepatitis C and human immunodeficiency virus in a real-world setting. Sci Rep. 2018;8(1):1858.

8. Condorelli F, Scalia G, Stivala A, et al. Detection of immunoglobulin G to measles virus, rubella virus, and mumps virus in serum samples and in microquantities of whole blood dried on filter paper. J Virol Methods. 1994;49(1):25–36.

9. Behets F, Kashamuka M, Pappaioanou M, et al. Stability of human immunodeficiency virus type 1 antibodies in whole blood dried on filter paper and stored under various tropical conditions in Kinshasa, Zaire. J Clin Microbiol. 1992;30(5):1179–1182.

10. Grüner N, Stambouli O, Ross RS. Dried blood spots--preparing and processing for use in immunoassays and in molecular techniques. J Vis Exp. 2015;(97).

11. Wrapp D, Wang N, Corbett KS, et al. Cryo-EM structure of the 2019-nCoV spike in the prefusion conformation. Science. 2020;367(6483):1260–1263.

12. Watanabe Y, Allen JD, Wrapp D, McLellan JS, Crispin M. Site-specific glycan analysis of the SARS-CoV-2 spike. Science. May 2020.

13. Faustini SE, Jossi SE, Perez-Toledo M, et al. Detection of antibodies to the SARS-CoV-2 spike glycoprotein in both serum and saliva enhances detection of infection. medRxiv. June 2020. doi:10.1101/2020.06.16.20133025

14. Jordan RE, Adab P, Cheng KK. Covid-19: risk factors for severe disease and death. BMJ. 2020;368:m1198.

15. Su X, Carlson BF, Wang X, et al. Dried blood spots: An evaluation of utility in the field. J Infect Public Health. 2018;11(3):373–376.

